# Traditional male circumcision and the risk for HIV transmission among men: a systematic review

**DOI:** 10.1101/2023.01.17.23284694

**Authors:** Gregorius Abanit Asa, Nelsensius Klau Fauk, Paul Russell Ward

## Abstract

**Introduction:** Although traditional male circumcision (TMC) is still practiced in a number of countries, and its healing process may have a high risk of HIV and other STIs transmission, there have been no published systematic reviews on TMC, HIV risk, and impacts on circumcised men and their families. The aim of this study is to synthesise evidence of how TMC practices contribute to HIV transmission among males and the impacts of HIV on themselves and their families.

**Methods:** The systematic search started with an initial search following the PICO (Population, Intervention, Comparison and Outcomes) framework. A systematic review was conducted to find literature using databases including PubMed, CINHAL, SCOPUS, ProQuest, Cochrane, and Medline. The search was limited to the English language, and with no year limit in order to capture as many articles as possible about circumcision, traditional male circumcision, HIV, and impact on men and their families. Critical appraisal tools developed by the Joanna Briggs Institute (JBI) for study design were used to assess the methodological quality of included studies.

**Result:** A total of 18 studies were included: 11 were qualitative studies, 5 were quantitative studies, and 2 were mixed-method studies. All the studies included were conducted in areas where traditional male circumcision was performed (17 in Africa and 1 in Papua New Guinea). The findings of the review were categorized into themes namely TMC as a cultural practice, consequences of not being traditionally circumcised on men and their families, and TMC-related risk of HIV transmission. The review showed that TMC and HIV risk could bring significant and negative challenges for men and their families.

**Conclusion:** The findings indicated the need for targeted health intervention programs and efforts to address psychological and social challenges in communities practicing TMC.

**Prospero Number Registration:** CRD42022357788.

## 1. Introduction

Circumcision is a cultural practice older than written history can explain, can be traced back to pre-Abrahamic times, and can be found in many Judeo-Christian traditions in Africa [1, 2]. It may be also one of the oldest human surgical procedures in the world [3]. It is a practice that has been widely performed on boys and young men by cutting off the foreskin of the penis as a rite of passage to mark the transition from childhood to manhood, primarily for religious and cultural reasons/beliefs [4, 5]. In many parts of the world, it has traditionally been practiced in Africa, Asia, Australia, Polynesia, and South and North America [3]. From the late 19^th^ century onwards, circumcision is not only seen as a cultural or religious practice/identity, but also a public health approach [6]. In the 1980s, observational studies came up with the hypothesis that circumcision might protect against Human Immunodeficiency Virus (HIV) transmission [7, 8].

Male circumcision has been identified to provide significant protection against HIV transmission and other sexually transmitted infections (STIs) in men [11-16]. This has been proven by Randomised Controlled Trials in South Africa, Kenya, and Uganda [15, 17, 18], showing that circumcised males were less likely to become infected with HIV. As a result, male circumcision is increasingly recommended as a strategy to reduce HIV transmission, particularly in areas of a high prevalence of HIV [19-28]. A report from the World Health Organization and the United Nations has also highlighted a correlation between the lack of male circumcision and higher HIV rates, specifically in Eastern and Southern Africa [29]. However, skepticism has also been raised regarding the protective effect of male circumcision on HIV transmission: some previous studies failed to prove the correlation between male circumcision and HIV infection prevention [30, 31], while some other studies found circumcision increased the risk of HIV transmission [32, 33]. Such skepticism seems to be supported by some evidence from Japan and Scandinavian countries showing that the percentage of circumcised men is low, but the prevalence of HIV cases in these counties is also low [34]. Furthermore, factors such as sexually active behaviors prior to circumcision, religion [35], history of STIs, and age [7] have been reported to be overlooked in the findings of randomised trials. These factors have also been as supporting reasons for doubt about the strength of the relationship between male circumcision and HIV transmission prevention.

Similar to medical circumcision, the protective benefits of traditional male circumcision (TMC) have been a common question. Some evidence has suggested that TMC provides less or no protection from HIV transmission due to less amount of foreskin removed [36-38]. Newly traditionally circumcised males are also considered to have minimal protection if they have sexual intercourse before the wound heals completely [15, 39]. The possibility of acquiring HIV infection through TMC is also considered high due to sharing of a surgical knife or blade on multiple men [24, 40-43]. TMC refers to the procedure of removing the foreskin on males in a non-clinical way by traditional circumcisers without formal medical training [44]. In addition to preparing newly circumcised males for the transition to manhood, TMC symbolises new initiates officially being accepted in the community with a new status of being a man and becoming a good model in family and society [45-47]. TMC also denotes that new initiates have a greater social responsibility to their families and community, act as negotiators in community disputes, and have a chance to learn about the community’s problems [19, 20]. These symbolisations highlight TMC as a sacred and secret rite. For example, in Africa, initiates are forbidden to talk with outsiders about the circumcision ritual and those who undergo the ritual as it will cause severe punishment imposed by the community [48, 49]. Similarly, sanctions will be imposed on females and non-circumcised males who gain information about the ritual [50]. To some extent, due to its sacredness, the further consequences of TMC practice have become a challenge for health intervention programs.

Studies on male circumcision and the risk for HIV transmission have been conducted in many parts of the world including low- and middle-income countries (LMICs) and developed countries. Although TMC is still practiced in a number of countries, and its healing process may have a high risk of HIV and other STIs transmission, to the authors’ knowledge, there have been no published systematic reviews on TMC, HIV risk, and impacts on circumcised men and their families. Thus, the authors consider it important to conduct a systematic review to synthesise evidence of how TMC practices contribute to HIV transmission among males and the impacts of HIV on themselves and their families. To determine whether a previous systematic review exploring this theme had been completed or is in progress, we conducted a preliminary search in PubMed, CINHAL, Scopus and in International Register of Systematic Reviews (PROSPERO) and found no underway systematic review on this topic in LMICs and developed countries. Therefore, this systematic review is needed to fill the gap and to help inform future health efforts at all levels including health practitioners, researchers, and policy makers.

## 2. Methods

### 2.1 The Systematic Search Strategy

The protocol for the systematic review has been registered with PROSPERO (registration ID: CRD42022357788) [51]. The systematic search started with an initial search following the PICO (Population, Intervention, Comparison and Outcomes) framework, which has been used as part of the WHO guidelines development process to inform evidence-based practice The systematic search was developed in collaboration with a health librarian expert, and the search terms were adjusted by each database. Databases searched included PubMed, CINHAL, SCOPUS, ProQuest, Cochrane, and Medline. The search was limited to the English language, and with no year limit in order to capture as many articles as possible about circumcision, traditional male circumcision, HIV, and impact on men and their families. The search strategies for the databases are in appendix 1. Medical Subject Headings (MeSH) were used as part of the search strategies. The search terms were formulated using the combination of key terms or the synonym of each concept using boolean terms (OR, AND). In addition to electronic search, Google scholar and google were used to search grey literature. Reference lists of all relevant articles were also scrutinised to identify articles that were not recaptured by electronic database search. The search was conducted 15 – 30 October 2022. The combination of key terms including the synonym of each concept is in table 1.

**Table 1.**
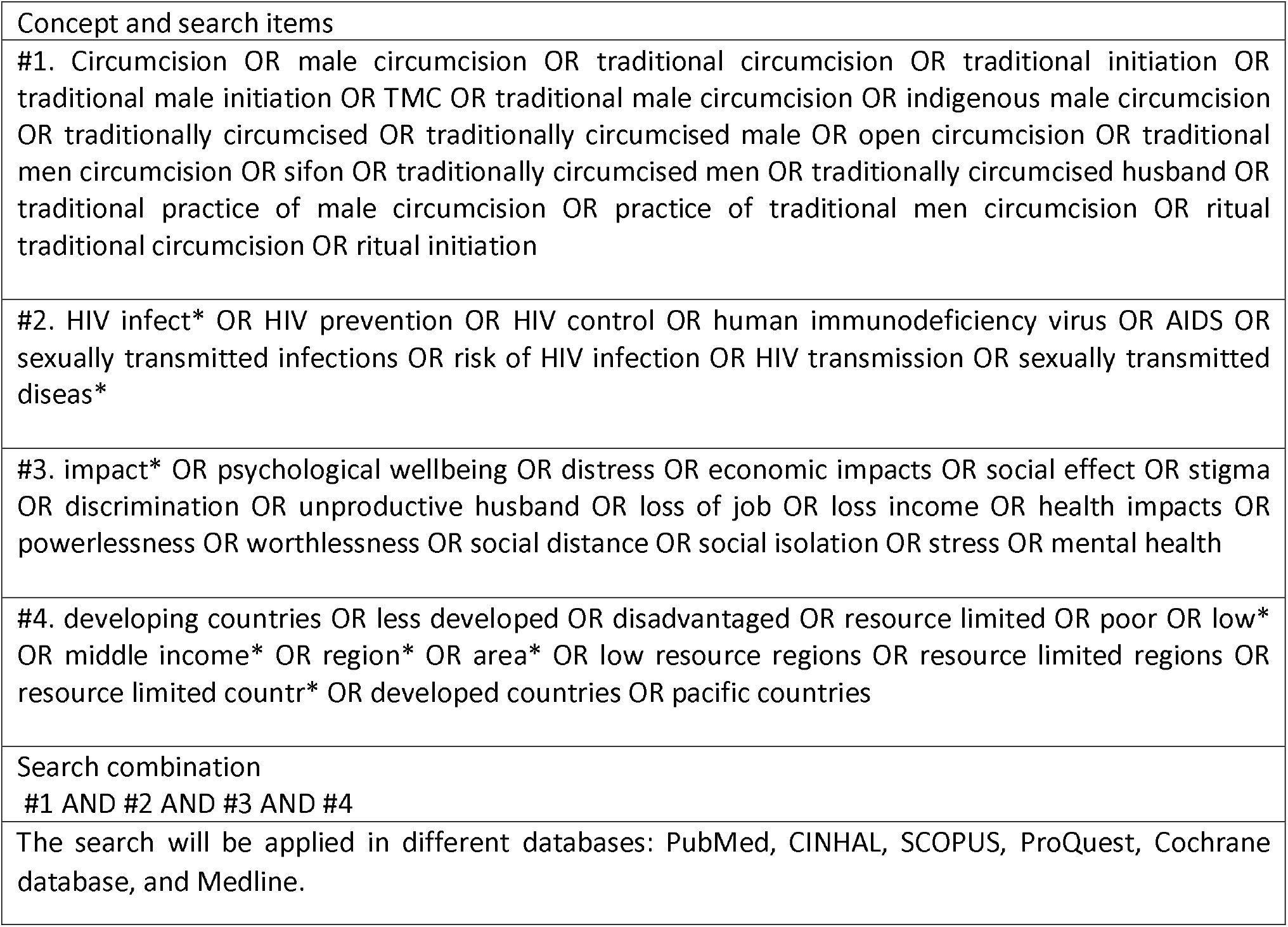
Search terms

### 2.2 Inclusion and Exclusion Criteria

The review included qualitative, quantitative and mixed method studies and evidence syntheses (systematic reviews). We also reviewed unpublished studies from reports and policy documents from google, google scholar, and WHO websites by applying relevant search terms. A summary of inclusion and exclusion criteria is shown in table 2.

**Table 2.**
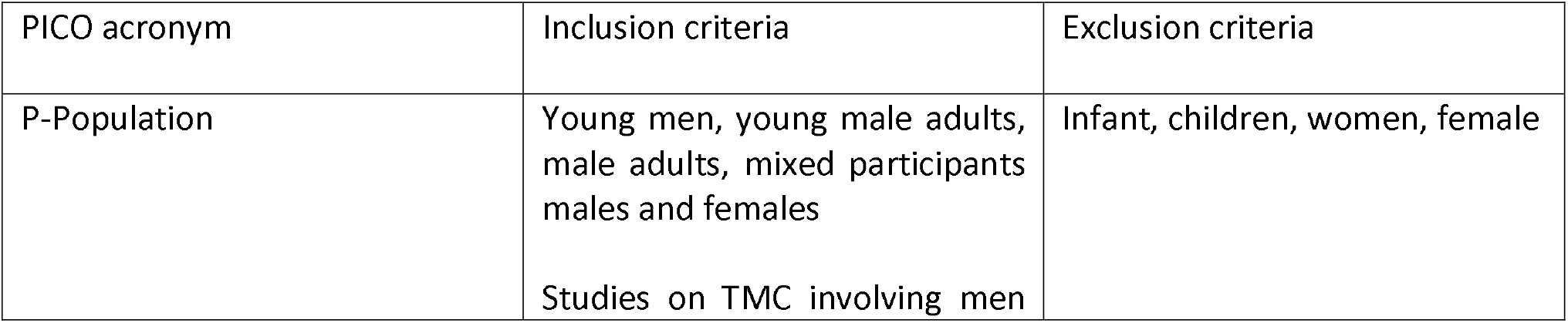
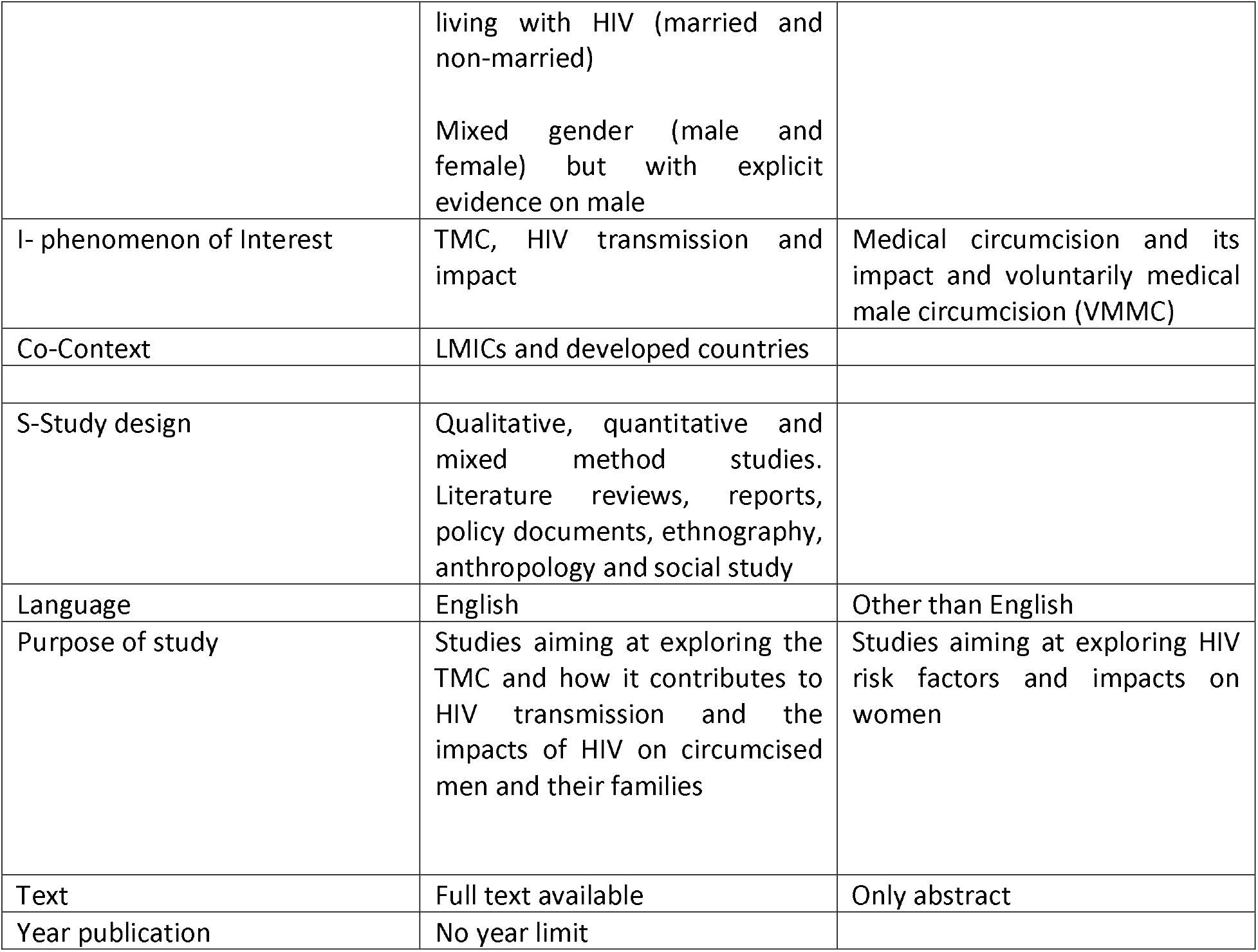
Inclusion and exclusion criteria

| PICO acronym | Inclusion criteria | Exclusion criteria |
| --- | --- | --- |
| P-Population | Young men, young male adults, male adults, mixed participants males and females<br>Studies on TMC involving men | Infant, children, women, female |
|  | living with HIV (married and non-married)<br>Mixed gender (male and female) but with explicit evidence on male |  |
| I- phenomenon of Interest | TMC, HIV transmission and impact | Medical circumcision and its impact and voluntarily medical male circumcision (VMMC) |
| Co-Context | LMICs and developed countries |  |
| S-Study design | Qualitative, quantitative and mixed method studies. Literature reviews, reports, policy documents, ethnography, anthropology and social study |  |
| Language | English | Other than English |
| Purpose of study | Studies aiming at exploring the TMC and how it contributes to HIV transmission and the impacts of HIV on circumcised men and their families | Studies aiming at exploring HIV risk factors and impacts on women |
| Text | Full text available | Only abstract |
| Year publication | No year limit |  |

### 2.3 Data Screening

All the identified articles (see Fig. 1) were collated and imported into EndNote X9 (Clarivate Analytics, PA, USA). The search identified a total of 3,041 articles. Duplicates (n=690) were removed using EndNote. The titles and abstracts of the remaining 2,359 articles were screened, further removing a total of 2,118 articles due to irrelevant populations and focus or aims. In total, 241 articles were examined in full text for eligibility. Of this, 222 articles were excluded due to not meeting inclusion criteria and one article not meeting methodological quality. After full text examination, 18 articles fulfilling inclusion criteria were finally included in the review. The 18 articles were then were assessed for methodological quality using critical appraisal tools developed by the Joanna Briggs Institute (JBI) for study design [56]. The methodological quality assessment was performed by the authors GAA and NKF. Uncertainty was resolved through discussion among the three authors. The screening process of the articles is reported and presented according to the Preferred Reporting Items for Systematic Review and Meta-Analysis (PRISMA) flow diagram (Figure 1) [57].

**Figure 1:**
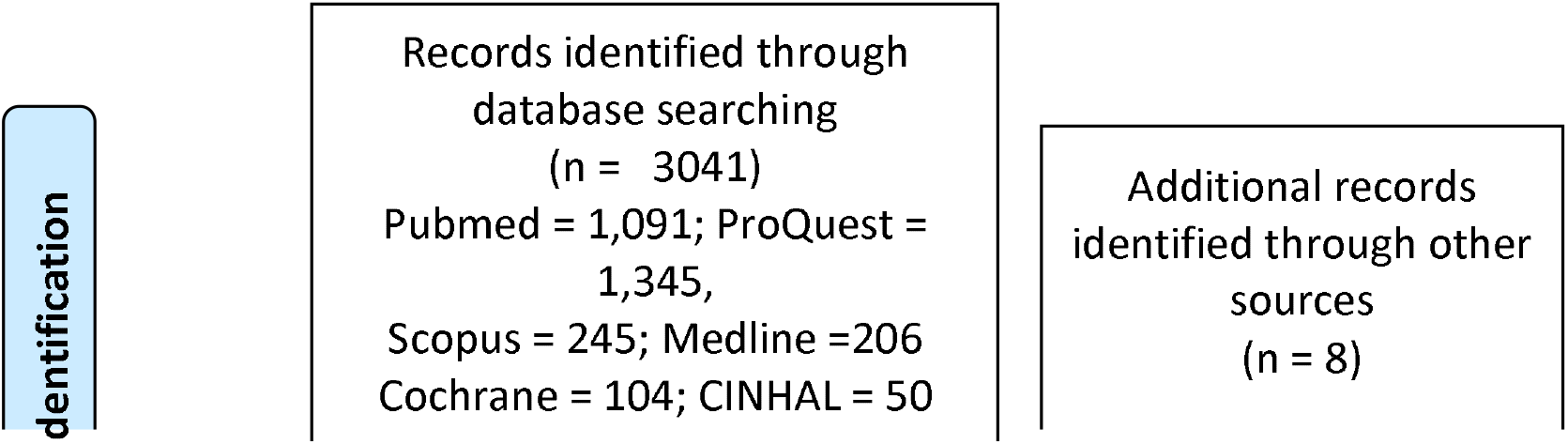
PRISMA Flow diagram of systematic literature search: records identified, removed, screened, and included in the review.

### 2.4 Data Extraction and Data Analysis

For each included article, data extraction was conducted with an extraction sheet. In the sheet, we recorded (i) study details: the last name of the first author, year of publication, study setting; (ii) study design: type of study, study aim, analysis methods; (iii) characteristics of participants: population, sex of participants, age of respondents; and (iv) results: the main themes, including TMC as a cultural practice, the impact of not being traditionally circumcised and the risk for HIV transmission (see Table 3). The analysis followed three-stage procedures by Thomas and Harden framework [58]: (i) coding the text line by line, interpreting the data, and identifying concepts or themes; (ii) developing descriptive themes by groping similar concepts in theme and sub-theme; and (iii) generating analytical themes by reviewing preliminary themes and discuss the addition or revision of the themes. The final analytical themes were then reviewed and decided as presented below.

## 3. Result

### 3.1 Characteristic of Included Studies

All included articles were published in English and were published from 2003 to 2020. The detail of the description of the studies is in Table 3. Among the 18 included publications, 11 were qualitative studies [39, 59-68], 5 were quantitative studies [69-73] and 2 were mixed methods [74, 75]. All the studies included were conducted in areas where traditional male circumcision was performed. A total of 48,468 participants were involved in the review, of whom 1055 and 47,413, respectively, were involved in qualitative and quantitative studies. 11 studies involved male only [62-67, 70-72, 74, 76], 7 studies involved men and women [39, 59-61, 69, 73, 75], 2 studies involved traditional circumcisers [61, 62], and 1 study involved health practitioners [61]. Participants’ ages ranged from 13 to 70 years old, however, 2 studies did not report the participant’s age [59, 63]. Most of the studies (n=17) were conducted in Africa while 1 study was conducted in Asia (Papua New Guinea) [59].

Key findings were grouped into three main themes, including (i) TMC as a cultural practice, (ii) TMC and challenges of not being traditionally circumcised on men and family, and (iii) TMC and the risk for HIV transmission. Finally, knowledge gaps were identified.

### 3.2 TMC As a Cultural Practice

It is widely recognized that TMC is practiced by various cultural groups among men as a rite of passage from childhood to adulthood. To the search, TMC is mostly practiced in LMICs in Africa and Asia. Thirteen studies [39, 59, 60, 62, 63, 65-68, 73-75, 77] discussed about TMC as a cultural practice: process of TMC, TMC as a secret and sacred practice, and reasons to undergo TMC.

#### 3.2.1 Process of TMC

Of the fourteen studies, seven studies [59, 60, 62, 63, 66, 67, 75] described three steps of TMC ceremony, including separation from family and community, transition, and incorporation into the family and community. In separation step, new initiates were taken to a mountain or camp for weeks or months [63, 75]. This long period was reported adequate time for healing process and learning about manhood [63]. The separation was meant for new initiates to demonstrate survival skills, such as ability to endure the pain which could improve men’s quality such as strength, courage, respect and fortitude [60].

Transition process is a step where initiates were taught about the social norms, cultural knowledge and community expectation for them so that they could socialize with their nuclear family, friends, and community [67]. For example, a study in Papua New Guinea [59] found that new initiates were taught about what they have as a clan, such as their ancestral values and spirit, their clan’s history, status, the land, the forest and the sea. Three studies [63, 66, 75] discussed about expectations in initiate’s families and communities after being traditionally circumcised which is in line with a study [78] reporting new initiates were expected to be a role model, have the ability to protect family, solve family disputes, and refuse tasks considered as a female domain. In the community, they were also expected to have sense of belonging to the community, take greater responsibilities (avoiding criminal activities and abuse of women), be able to cooperate with elders, and have the ability to face difficulties in the future.

In addition to learning about family and community, several studies [67, 75] reported that new initiates were taught about sexuality during TMC ceremony. A study in Limpopo, South Africa [60] found that sexual socialization during TMC puts an emphasis on sexual control and sexual reserve rather than “permit to sex.” For example, initiates were taught that if they did not wait long time to have sexual intercourse after being circumcised, their foreskin will grow again, and therefore, they have to undergo a new circumcision which is more painful [67]. However, other findings [67, 75] discovered that the emphasis of sexuality during circumcision has been changed with circumcision as a “license” for sex including unsafe sex behaviors. These studies support the findings of another study reporting that traditionally circumcised men tended to assume that they had unlimited and unquestionable rights to have access to sex [78].

Incorporation process was marked by the return of initiates to the family and community. In South Africa [67], upon returning new initiates wore a new dress code symbolizing new circumcised men reentering family and community as a new individual or a transformed individual who were ready to fulfill new roles in their society. This process is marked with celebration by slaughtering animals (a goat or a sheep) as a sign of thanks to ancestors, family and community [63]. A study in Papua New Guinea found that incorporation was marked with having a celebration or party with family and community [59]. Celebration of successful traditional circumcision draw symbolic power of being custodians of cultural practices resulting in a sense of community, social identity, and belonging [62].

Three studies [40, 62, 70] described TMC as an incomplete or partial circumcision where not all foreskins were removed. This is usually performed in non-clinical settings by traditional circumcisers without formal medical training. The left foreskin is considered the same as not being circumcised as the foreskin keeps semen in the penis, thus, making them “dirty” and vulnerable to easily being infected with HIV and other STIs infections compared to full circumcision (medical circumcision) [62]. Findings showed that TMC, similar to medical circumcision, may reduce the risk of HIV and other STIs, however, the amount of foreskin removed during the ceremony determines the extent of effectiveness against HIV transmission.

#### 3.2.2 TMC As a Secret and Sacred Practice

Six studies [59, 60, 62, 63, 67, 73] described TMC as a sacred, secret, and compulsory cultural practice in communities. As a sacred and secret practice, TMC was conducted with certain rituals in certain places and performed my certain people (traditional circumcisers). In Tanzania, the traditional circumcisers were appointed by ancestors through dreams, and the skills were passed from one person to another through observation [62]. Meanwhile, in Xhosa, South Africa, the skills were taught by elder circumcisers through apprenticeship [63]. Ritual ceremony was performed by traditional circumcisers or clan leaders prior to circumcision. Similarly, as a compulsory practice, all men within community were required to undergo such practice. Secretness is also marked by separation or isolation. Studies in Africa found that secretness is marked by isolating or separating new initiates from their families and communities [63, 67]. Similarly, a study in Papua New Guinea [59] found that TMC was performed in a designated home for exclusive use of men where only men were allowed to witness the actual process.

The cultural practice of TMC in Africa and Asia does not allow women to be around the ceremony and view or have knowledge of the process of TMC. It is believed initiates will be affected by witchcraft and experience slow recovery process if women were present during the ceremony. However, women in Papua New Guinea [59] were found highly knowledgeable about the whole process of TMC, able to explain in detail the cutting process, the procedures and the disposal of blood. The role of women in the community in Papua New Guinea was to start preparing for welcoming new initiates such as making food, buying pigs to be eaten during celebration, singing, dancing and giving gifts.

The sacredness of the TMC was reported to be related with the initiate’s ancestors intervention as highlighted in two studies [62, 63]. In South Africa, ancestors were reported to be involved in TMC process and wound healing following circumcision. Long healing wounds or not healing properly is associated with sexual impurity. For example, in Monduli, Tanzania [73], it was believed that the wound took two weeks to be completely healed for initiates who had not engaged in sexual intercourse prior to circumcision, and took more than one month for the exposed ones. Due to this, in certain communities, initiates were asked to repent their sins so that the wound heals quickly [63].

#### 3.2.3 Reasons to Undergo TMC

Ten studies [39, 59, 62, 63, 65-68, 73, 74] describe rationales for TMC. These studies underlined an obligation for performing cultural rites to prepare new initiates for the responsibility of adulthood as the main reason for TMC. A qualitative study in South Africa [39] found that men and women underlined the importance of TMC to live up to cultural values and community expectations. They believed that traditionally circumcised men were more mature, less abusive, and more responsible, compared to non-traditional circumcision as they had received teachings during ceremonies. Furthermore, learning social norms, cultural values and men’s related values such as being tough and brave to take risk were aspects that were only found in traditional circumcision and not in medical circumcision [79]. This reason seemed to significantly influence initiates’ resistance to the modern medical circumcision. Expecting the privilege of being accepted and being together, such as having meals in the same dishes with the circumcised ones, was also a supporting factor for men to undergo TMC [74].

Four studies [73, 77, 80, 81] described about economic reasons to undergo TMC. Low cost for TMC compared to medical circumcision was reported to affect initiate’s and their family’s decision [80]. Evidence from South Africa showed that new initiates could not afford to pay medical circumcision and the amount of money charged by legal traditional circumcisers resulting in new initiates took health risk visiting illegal traditional circumcision charging lower price [74]. Such evidence seemed to show that people who were economically vulnerable in traditional settings may only be able to access cheaper circumcision services with high risk of complication and potential risk of HIV transmission. Nevertheless, in many cases, the cost charged for traditional circumcision did not include the time the wound was fully recovered, complication requiring further medical treatment, and celebration of fully recovery [77].

Five studies [39, 59, 62, 66, 68] discussed the influence of women (e.g., girlfriends, future wives/partners), family, community and peers on men to undergo circumcision (TMC and medical circumcision). Evidence from South Africa showed that women often scheduled the appointment for their boyfriend or and husband to be traditionally circumcised [76]. Similarly, another finding in South Africa showed that women tended to undermine the manhood of non-circumcised males [66]. Also, a finding in PNG showed that women prefer circumcised men for marriage and as a sexual partner [59]. In addition to cultural reasons, women’s preferences for circumcised men were related with pleasure and satisfaction during sexual intercourse compared to uncircumcised men [39]. Family, community and peers were also reported as significant influences for young men to undergo TMC [62].

### 3.3 TMC and The Consequences of Not Being Traditionally Circumcised on Men and Their Families

Eleven studies [39, 59-62, 64, 65, 67, 69, 73, 82] described the challenges of not being traditionally circumcised, including psychological impacts and social challenges. The details about these aspects are presented below.

#### 3.3.1 Psychological Challenges

Psychological impacts including feelings of shame, stress, and embarrassment were common negative challenges experienced by men who were not traditionally circumcised [83]. Such challenges were supported by experiences of being asked by friends about when to undergo TMC [83]. Another stressor for such psychological challenges included feeling obligated to undergo the ritual. Similarly, uncircumcised men were negatively affected by community perception on masculinity and adulthood.

Social pressures associated with traditional circumcision was another stressor for psychological challenges facing young people in some settings. Several studies described about adolescents and young men in Africa who experienced social pressure from their family and peers for being medically circumcised and uncircumcised [39, 62]. For example, a number of men acknowledged that they decided to be traditionally circumcised because their fathers or brothers had undergone circumcision, leading them to feel obligated to undergo the same ritual [67]. Others pointed to the respect to culture or system they grew up with where all men underwent the same ritual [67]. In Xhosa community, South Africa, it was often uncircumcised men were called cowards by friends at the same age [67]. Therefore, the decision to be traditionally circumcised was to avoid being harassed and ridiculed. In the family context, pressure of young men to be traditionally circumcised stems from the desire to maintain the family honor [64].

Another significant pressure was from women. Studies found that boys felt pressure when asked by girl friends or partners about their circumcision status. A study in South Africa found that girls were considered trivial if dating and walking with uncircumcised boys [67]. They were also considered as not ready for building a relationship with women [64]. Another finding in Africa also showed that circumcision is beneficial for women who were married to men who were cheating as circumcision might protect against HIV transmission [67].

#### 3.3.2 Social Challenges: Stigma, Discrimination and Disrespect

Seven studies [39, 59, 60, 64, 66, 67, 82] described stigma and discrimination related to TMC. A study in Xhosa, South Africa noted that 70% of Xhosa initiates felt that they would experience stigmatization if they were not traditionally circumcised [84]. In the same study setting, uncircumcised men and those underwent medical circumcision were stigmatized as boys who were immature and impossible to distinguish them from ‘real men’ [64]. Similarly, uncircumcised men in PNG [59] felt ridiculed, mocked and people made fun of those who were not traditionally circumcised. Indeed, uncircumcised men in PNG are referred to as utilusa (foreskin) instead of using their actual name. Such impact was not only experienced by the initiates but also the initiate’s families in which the initiates’ father and family were looked down by others within the community. For young uncircumcised men in Africa, stigma, discrimination, and rejection were reported to have caused long-term psychological effects reflected in anxiety, personality change and lack of confidence [64].

It is also reported that uncircumcised men were treated differently and assumed negatively as reported in two studies [64, 67]. In the family and community, they were highly vulnerable, often blamed for any inappropriate actions and considered incapable of moral worth. For example, uncircumcised men are often accused of being liars and thieves and were also treated like animal (a dog) in their community [64]. Another study in Africa showed that uncircumcised men and those underwent medical circumcision would not be accepted in the community, not obtain rights and responsibility in their family, and had no rights to negotiate with elders [67]. Also, they are not allowed to start families within their community and are not allowed to inherit and have property on their own [64]. Such negative impacts were reported to affect uncircumcised men psychologically, such as feeling embarrassed, disadvantaged and having low/no moral worth.

A couple of studies also suggested that men who were uncircumcised and underwent medical circumcision did not earn respect from community [39, 67]. In some settings it is considered proper for the community not to respect men who failed to follow the rite of passage and this leads them to not receive the same status as other men [39, 67]. Uncircumcised men and those who failed to follow the ritual would be marginalized from traditional ceremony and community discussion [64]. These studies suggested that such consequence can lead to further psychological problems such as feeling sad, low self-esteem, feeling guilt, social withdrawal and frustration among traditionally uncircumcised men.

The social challenges, stigma, discrimination and expectation towards traditionally circumcised men underline cultural constructions of the penis and body which then leads to construction of masculinity and womanhood, which further raises issues of gender constructions [85]. The body functions metaphorically symbolize social status, tribal affiliation, family position, and gender [85]. Rite of passage indicated by ritual and social transformation plays significant roles in social interaction within community [85].

### 3.4 TMC and the Risk for HIV Transmission

Nine studies [39, 60-62, 69, 70, 72, 73, 75] described about (i) shared knife and bandage, unhygienic environment and the risk for HIV transmission; (ii) TMC promoted multiple sexual intercourse and increase sex partners, (iii) Belief in the protective effects of TMC against HIV/AIDS, and (iv) TMC and Knowledge of HIV Transmission.

#### 3.4.1 Shared a Knife and Bandage, Unhygienic Environment and the Risk for HIV Transmission

Four studies [61, 62, 69, 73] highlighted the practice of one knife or blade used to circumcise several initiates. For example, the majority of participants in a study in Tanzania reported that one knife was used in all TMC ceremony [62]. Using one knife or blade to circumcise several initiates in one or several TMC ceremonies were reported to put initiates at high risk of being infected with HIV and other STIs as some of the initiates may have had unsafe sexual intercourse prior to circumcision and may already be HIV-positive [69]. However, another finding [73] showed that some traditional circumcisers started using one knife or razor one for one initiate.

A study by Mpateni and Kang’ethe [61] also highlight the possibility of being infected with HIV and other infectious diseases through sharing bandage and unhygienic environment reflected in unclean areas around the ceremony and using unwashed dishes to eat. Such poor environment was supported by careless mistakes of traditional circumcisers who lack of knowledge of the importance of hygiene and the way the infectious diseases spread.

#### 3.4.2 TMC Promotes Multiple Sexual Intercourses and Increases Sex Partners

Promoting multiple sexual intercourse in TMC was reported in five studies [39, 60, 61, 72, 75]. A qualitative study in Malawi [75] found stakeholders’ concern on the role of TMC ceremony promoting sexual adventure among new initiates, asserting that circumcised men were not children anymore after they had sexual intercourse following circumcision. Similarly, there was also myths and false teaching that after being traditionally circumcised, initiates had to have sex with several females for testing of the penis [61]. As a result, many boys took this ceremony as a license to start having sex. This finding supports the finding of a study [60] that traditional initiation school had a strong influence on initiates sexual behaviors. This high sexual desire was reported to be supported by considerable amount of time they spent in the bush or camp during TMC ceremony without any contact with female [39]. Elsewhere, a qualitative study [39] found that traditionally circumcised men were told to have sexual intercourse without condoms to prove that they could enjoy flesh-to-flesh sex following the circumcision. As a result, some initiates continued to not using condoms following TMC.

Promoting sexual intercourse have led traditional initiates to increase the number of sex partners as reported in two studies [70, 72]. The study in Kenya found some initiates had more sexual desire following TMC, resulting in initiates increased their number of sexual partners. Such practice was reported to increase the transmission of STIs [72]. The study suggests the need of the synergy between traditional ritual and medical intervention for HIV preventive practice.

#### 3.4.3 Belief in the Protective Effects of TMC Against HIV and Condom Use

Belief in the protective effects of TMC against HIV/AIDS transmission was also a risk factor which further affects initiates’ sexual behaviors. Four studies [67, 71, 72, 86] discussed about beliefs in the protective effects of TMC. Traditionally circumcised men tended to believe that TMC offers complete protection against HIV and other STIs and that circumcision is an alternative of condom use [86]. A quantitative study in Eastern Cape, South Africa found that 97% of TMC initiates believed that TMC made initiates become a ‘real man’, and that they did not need to use condoms during sexual intercourse [72]. A study in Sub-Saharan African countries [70] found that traditionally circumcised males were less likely to use condoms following circumcision. This is similar with the findings from Eastern Cape [71], reporting TMM initiates were more likely to engage in risky sexual activities. Similarly, a cohort study in South Africa [72] found that 38% of traditionally circumcised men reported inconsistent condom use when having sex, and 8% of them reported never using condoms.

#### 3.4.4 TMC and Knowledge of HIV Transmission

Lack of knowledge of HIV and other STIs among initiates and traditional circumcisers were reported in five studies [60-62, 70, 72]. Similar to medical circumcision, TMC initiates also believed that TMC protected them from STIs such as syphilis and gonorrhea and enhances personal hygiene [62]. A cohort study [72] found that new initiates who went through traditional circumcision were mainly for cultural reasons, rather than HIV prevention. Absence of information about HIV and other STIs prior to and after the circumcision was also reported as a HIV risk factor. For example, a study in Limpopo [60] found that traditional initiation school did not provide information about sexual health and HIV/AIDS and other STIs but tended to encourage new initiates to engage in risky sexual activities. Safer sexual behavior such as condom use and being faithful with one sex partner was not considered a part of initiation school programs. This was acknowledged by initiates, who said that they obtain the information about condom from local clinics and mass media [60]. A qualitative study in South Africa [67] found that absence of information has led to lack of understanding about the correlation between circumcision and HIV transmission.

Lack of knowledge of the mode HIV transmission was not only in TMC initiates but also among traditional circumcisers reflected in encouraging sex adventure, using one knife for several initiates, sharing bandages for several initiates, and ignorance of unhygienic environment [61]. A study in Tanzania [73] revealed that most of the traditional circumcisers did not associate between traditional circumcision practice and HIV/AIDS, assuming that HIV/AIDS was an urban disease. However, another finding of the same study also showed that careless mistakes performed by traditional circumcisers by not using any protection such as gloves when cutting the foreskin of the penis increased the risk of HIV transmission.

## 4. Discussion

### 4.1 TMC Practices and HIV Transmission

The findings show evidence that TMC as a cultural practice remains practiced in some communities in LMICs in Africa and Asia. The majority of the studies [39, 59-63, 65-68, 73-75, 77] reported that TMC in communities is not merely to cut off the foreskin but also to live up the tradition, keep the relation with their ancestors, and to teach and inherit cultural values and the values of ‘manhood’ to new initiates. The practice of TMC is highly valued as a secret and sacred practice, taking weeks and months from the separation step until the new initiates returned to the families and communities. Secretness and sacredness aspects in TMC may have led to difficulties to health intervention to control safety procedure. Such practice and its potential health risk factors reflects the high value the community puts on culture or tradition rather than any other types of medical or modern health intervention.

Studies in many communities in Africa found that TMC is a compulsory practice where all men were required to be traditionally circumcised, leaving challenges at individual and family level for those who did not undergo such practice. At the individual level, TMC causes psychological impacts for uncircumcised men and those followed medical circumcision including feeling ashamed, stressed, and pressured. These impacts were supported by the cultural values that put TMC as a standard of maturity of men. In addition to experiencing pressure from family and community, uncircumcised men also felt pressure from girls or women who preferred to build a relationship or to have sexual intercourse with traditionally circumcised men [39, 59, 62, 66]. Such impacts were also attributed for those who were not completely follow the process of TMC or mixed with medical circumcision. Although studies included in this review did not report the challenges of TMC on family, it is plausible to argue that family would be impacted if young men within the family did not undergo TMC.

Not undergoing TMC could also lead to negative social challenges such as stigma, discrimination, and disrespect towards men [60, 64, 82]. For example, those who did not undergo TMC could be labelled immature, irresponsible and easily ridiculed, humiliated, and mocked. In families and communities, traditionally uncircumcised men were stigmatized as the cause of any crime or irresponsible actions. Similarly, they did not have full rights to talk, discuss and negotiate with elders about families and communities’ problems. They are labelled and treated without a respect (e.g., like a dog) which implies that they are considered less than human. Such impacts are in line with the components of stigma, such as labeling human differences, hegemony cultural practice associated labelled persons to undesirable characteristics, labeled persons are separated with the term “us” and “them”, labelled persons experience loss of status and discrimination, and labelled persons experience difficulties in access to social, economic and political power [64, 87]. Similar to psychological impacts, all the studies included in the review mostly focus on stigma on initiates and thus less concern on stigma on family. Stigma, discrimination, and disrespect experienced by initiates prior to circumcision and uncircumcised men also reflect lack of social and psychological support from their families, friends, and communities.

TMC is generally assumed to have implications for HIV transmission [39, 60, 61, 69, 70, 72, 73, 75]. Unsafe procedure of TMC practices such as using one knife to circumcise several initiates, not wearing gloves when circumcise initiates, and unhygienic environment, raising the concern of on potential spread of infectious diseases, including HIV [61, 69, 73]. In addition to learn about culture and manhood in the transition period, initiates were also taught about exploring their sexuality, leading initiates to consider TMC as a ‘gateway’ to have unquestionable sex adventure and to have more than one sexual partner. For example, initiates were asked to have sexual intercourse with women who have had sex before as reported in a previous study. For example, initiates were asked to have sexual intercourse with women who have had sex before which is in line with another study [88] reporting that initiates were required to have sexual intercourse without protection several days before the wound heals as a way to speed up the recovery process The correlation between TMC and the risk of HIV transmission is also related with the belief that TMC has the same protective effects as using a condom. This belief may also be supported by the sacredness aspect of TMC rite, believing that the dead ancestors will intervene in the health of the initiates as in line with previous studies [59, 73]. Another supporting factor for TMC and the risk of HIV transmission is lack of knowledge on the mode of HIV transmission. In some communities, safe sexual behavior was not part of the subjects taught during TMC rite, leading initiates to have no knowledge about HIV risk. This is in line with a finding in another study among 100 participants, of whom 67% of them were not aware of the risk of traditional circumcision for HIV transmission [89]. However, the risks for HIV transmission were also reported among initiates who had knowledge about HIV transmission. Findings of a previous study suggest that circumcised men who had knowledge about HIV preventive measure of male circumcision and believed that male circumcision could reduce the risk of HIV infection were more likely to engage in risky sexual behaviors or sex without condoms with multiple partners [90]. The risks for HIV transmission in the practice of TMC reflect lack of education, public awareness campaign and counseling for young men, parents, students, local leaders, and traditional circumcisers in the community practicing TMC.

### 4.2 Implications for Future Intervention

The systematic review provides a range of negative impacts of not being traditionally circumcised on men and scant information on the impacts on their families. Overall, the studies highlight psychological and social challenges that need to be addressed in communities practicing TMC. The studies also highlight TMC and the risk for HIV transmission which require future health interventions.

In this review, it is obvious that stigma, discrimination, and disrespect towards uncircumcised men or those who followed medical circumcision were within initiates’ family and communities. This is because TMC is viewed more prestigious than any other circumcisions. It is suggested to have continuous counseling, approach, and education on communities where traditional beliefs and norms are still highly valued [60]. These approaches should reach not only family but also community and school. In light of the TMC and the risk for HIV transmission, it is noted that in some communities TMC has no role to play in preventing HIV and other STIs transmission such as promoting multiple sexual intercourse, not using condoms, and believing the full protection of circumcision against HIV transmission. To address this problem, education to target traditional circumcisers, traditional leaders, parents, and young men are required in order to improve the safe practice and prevent HIV transmission as reported in several studies [60, 77, 91]. Similarly, education on condom use and free, accessible condoms should also reach the camps where TMC practices were performed [60]. In addition, service delivery on providing free HIV testing for initiates in communities practicing TMC is needed.

### 4.3 Strengths and Limitations of the Study

Although many studies on male circumcision have been conducted mostly in Africa and some in Asia, this review is, as far as the researchers know, the first known study on TMC, the risk for HIV transmission and impacts on them and their families. The use of six databases and multiple search terms across 18 included studies helped the researchers conducted a comprehensive systematic review of the literature and provided a broad range of studies in LMICs and developed countries. The inclusion of qualitative, quantitative, mixed methods helps the researchers to collate the current knowledge and knowledge gaps aimed the risk factors and impact of TMC on men and their families. Finally, the publications, the study selection methods, and the appraisal process altogether provided a substantial evidence that supports the key findings reported in the literature review. However, the literature review only included articles published in English which may have narrowed the scope and the authors may have missed the topic reported in other languages.

### 4.4 Implications for Future Studies

The review of the literature documents existing evidence and knowledge gaps about TMC, HIV risk, and its impact on men and their families. The review of the literature suggests that the previous studies mainly focus on the correlation on TMC and the risk for HIV transmission; none has explored TMC, HIV risk and its impacts on men and their families and none involved traditionally circumcised men living with HIV. Similarly, most of the included studies were in Africa settings, only one study was in Asia. Exploring TMC practice in different settings other than in Africa can help understand the similarities and differences of TMC practices and the implicaion on HIV transmission and its impact on men and their families. The review found very limited studies involved wives of married men who have done traditional circumcision and women that have unprotected sexual intercourse with newly traditional circumcised men to explore their views and sexual practices in relation to TMC. Furthermore, none of the included studies explored the views of health professionals and policy makers on TMC and its possible negative health consequences and how these have been addressed at policy level. Also, there is very limited studies exploring traditional circumcisers’ views on the TMC and HIV risk. Future studies are required to fill these gaps of knowledge which may provide useful information for the development of specific interventions for safer TMC and preventing HIV and other STIs transmission.

## 5. Conclusion

The review presents three main themes namely TMC as a cultural practice, consequences of not being traditionally circumcised, and TMC-related risk of HIV transmission. These themes provide evidence that TMC and HIV risk could bring significant and negative challenges for men and their families. This review may be useful in designing programs to address social and psychological impacts associated with TMC practice in communities and integration of health intervention with medical circumcision.

## Data Availability

All data produced in the present work are contained in the manuscript

https://www.crd.york.ac.uk/PROSPERO/#searchadvanced

## Contributors

Conceptualization and the development of the protocol, Gregorius Abanit Asa (GAA), Nelsensius Klau Fauk (NKF), and Paul Russell Ward (PRW); Methodology, GAA, NKF and PRW; systematic search of literature, GAA and NKF; formal analysis, GAA; writing-original draft preparation, GAA; writing-review and editing, GAA, NKF, and PRW; supervision, GAA, NKF, and PRW. All authors have read and agreed to the published version of the manuscript.

## Patient and public involvement

Patients and/or the public were not involved in the design, or conduct, or reporting, or dissemination plans of this research.

## Patient consent for publication

Not applicable.

## Ethics approval

Not applicable.

## Provenance and peer review

Not commissioned; externally peer reviewed.

## Data availability statement

All data generated or analysed during this study or review are included in this published article.

## Supplemental material

This content has been supplied by the author(s). It has not been vetted by BMJ Publishing Group Limited (BMJ) and may not have been peer-reviewed. Any opinions or recommendations discussed are solely those of the author(s) and are not endorsed by BMJ. BMJ disclaims all liability and responsibility arising from any reliance placed on the content. Where the content includes any translated material, BMJ does not warrant the accuracy and reliability of the translations (including but not limited to local regulations, clinical guidelines, terminology, drug names and drug dosages), and is not responsible for any error and/or omissions arising from translation and adaptation or otherwise.

